# SARS-CoV-2 Acquisition and Immune Pathogenesis Among School-Aged Learners in Four Diverse Schools

**DOI:** 10.1101/2021.03.20.21254035

**Authors:** Dan M. Cooper, Michael Z. Zulu, Allen Jankeel, Izabela Coimbra Ibraim, Jessica Ardo, Kirsten Kasper, Diana Stephens, Andria Meyer, Annamarie Stehli, Curt Condon, Mary E. Londoño, Casey M. Schreiber, Nanette V. Lopez, Ricky L. Camplain, Michael Weiss, Charles Golden, Shlomit Aizik, Bernadette Boden-Albala, Clayton Chau, Ilhem Messaoudi, Erlinda R. Ulloa

## Abstract

**Background:** Understanding SARS-CoV-2 infection in children is necessary to reopen schools safely.

**Methods:** We measured SARS-CoV-2 infection in 320 learners [10.5 ± 2.1(sd); 7-17 y.o.] at four diverse schools with either remote or on-site learning. Schools A and B served low-income Hispanic learners; school C served many special-needs learners; and all provided predominantly remote instruction. School D served middle- and upper-income learners, with predominantly on-site instruction. Testing occurred in the fall (2020), and 6-8 weeks later during the fall-winter surge (notable for a tenfold increase in COVID-19 cases). Immune responses and mitigation fidelity were also measured.

**Results:** We found SARS-CoV-2 infections in 17 learners only during the surge. School A (97% remote learners) had the highest infection (10/70, 14.3%, p<0.01) and IgG positivity rates (13/66, 19.7%). School D (93% on-site learners) had the lowest infection and IgG positivity rates (1/63, 1.6%). Mitigation compliance [physical distancing (mean 87.4%) and face covering (91.3%)] was remarkably high at all schools. Documented SARS-CoV-2-infected learners had neutralizing antibodies (94.7%), robust IFN-γ+ T cell responses, and reduced monocytes.

**Conclusion:** Schools can implement successful mitigation strategies across a wide range of student diversity. Despite asymptomatic to mild SARS-CoV-2 infection, children generate robust humoral and cellular immune responses.

**Key Points:**

- Successful COVID-19 mitigation was implemented across a diverse range of schools.
- School-associated SARS-CoV-2 infections reflect regional rates rather than remote or on-site learning.
- Seropositive school-aged children with asymptomatic to mild SARS-CoV-2 infections generate robust humoral and cellular immunity.

## INTRODUCTION

An urgent need for data on SARS-CoV-2 incidence, immune mechanisms, and mitigation fidelity in the unique setting of K-12 schools was recognized at the earliest stages of the COVID-19 pandemic^1^ when K-12 schools closed in the U.S. and across the world. In this report, we summarize the results of the *Healthy School Restart Study*, a prospective study of four diverse schools in Orange County, California, at two distinct phases of the COVID-19 pandemic: 1) early in the fall (2020) school semester, at a relatively low level of community COVID-19 case rates of approximately 3-4 cases per 100,000 across the county (in September 2020, surveillance rates for the county were estimated at 12% but 17% in communities of color^2^), and 2) approximately 6-8 weeks later in the midst of the fall-winter surge in which COVID-19 had increased to about 40 cases per 100,000. We tested the assumption used to support school closures that learners would be less susceptible to viral infection if they avoided on-site learning^3–5^. Key objectives were 1) to begin to understand SARS-CoV-2 infection in schools that reflected the diversity of our region, 2) to gain insight into the serological and cellular mechanisms in the pediatric population in response to SARS-CoV-2 infection, and 3) to measure the fidelity of SARS-CoV-2 mitigation procedures.

## METHODS

### Design

A total of 320 learners [mean age 10.5 ± 2.1 (sd); range 7-17 y.o.] and 99 school staff enrolled in our study across four schools for two testing cycles. Participants were allowed to enroll in the study at the second cycle even if they did not participate in the first visit. During the first cycle, 181 students aged 10-13 y.o. were enrolled. During the second cycle, 161 learners returned, and 139 new learners aged 7-17 y.o. were enrolled to accommodate additional requests for testing by the schools. During the first cycle, 99 adults were enrolled, and during the second cycle, 90 returned. At each of the testing cycles, each participant underwent:

1. Brief medical history and a COVID-19 symptom screening
2. Anterior nasal swab for SARS-CoV-2 and co-circulating respiratory pathogens
3. Optional non-fasting phlebotomy for serological and other immunological markers of SARS-CoV-2 infection

Pediatric participants were also offered a non-fasting lipid screening as an added benefit to the optional phlebotomy at cycle 2, as this screening test is recommended by the American Academy of Pediatrics.

### School Selection and Study Participants

We partnered with four schools that reflected the diverse population of Orange County ensuring adequate representation of low-income, minority, and special-needs learner-participants (Table). Inclusion criteria for the student participants were age (7-17 y.o.), current enrollment at one of the schools participating in the study, and fluency and literacy in English or Spanish. The criteria for adult school staff participants were age (equal or greater than 18 years), current employment at one of the participating schools, and fluency and literacy in English or Spanish.

### IRB Approval and Consent

The study was approved by the institutional review boards at the Children’s Hospital of Orange County (CHOC) and the University of California Irvine (UCI). Informed assent from the children and informed consent from parents or legally authorized guardians, or from the adult participants, were obtained remotely or in person.

### Approach to Participation

We organized virtual meetings with school staff and teachers to explain the study and to answer questions at each school. We also held town-hall-type meetings with potential students and their parents or authorized legal guardians. Meetings were held in both English and Spanish. We positioned our testing setup out of the way of normal school operations.

### Identification of SARS-CoV-2 and Co-Circulating Respiratory Pathogens

Anterior nasal swabs were obtained from each participant. The BIOFIRE Respiratory 2.1 Panel was used to identify SARS-CoV-2 in addition to 21 other respiratory pathogens by RT-qPCR done in the CHOC Clinical Laboratory. Per state mandates, positive SARS-CoV-2 findings were reported by the laboratory to the Orange County Health Care Agency (OCHCA) for subsequent confidential action and tracing by county health authorities.

### Blood Samples for Immune Response and Lipid Profile

Whole blood samples were collected in CPT tubes from 274 of the 320 learners. Plasma and peripheral blood mononuclear cells (PBMCs) were isolated by centrifugation. Presence of nucleocapsid protein (NP)-specific IgG was determined with the Abbott Architect immunoassay. IgM and IgG antibody titers against the NP and the receptor binding domain (RBD) of the spike protein were measured using standard ELISA^6^. Specific SARS-CoV-2 neutralizing antibodies were measured using focus reduction neutralizing assays^6^. IFN-γ producing T cells following stimulation with overlapping peptide pools were determined using mAB ELISpot plates and PBMCs from 34 SARS-CoV-2 seropositive samples and 34 age- and sex-matched uninfected controls from enrolled participants^7^. Circulating immune and inflammatory mediators (e.g., TNF-α, IL-6) were also measured using the Human 45-Plex kit from R&D^7^. Immunophenotyping to identify innate and adaptive cells was done using flow cytometry in 15 SARS-CoV-2 seropositive samples and 15 age- and sex-matched controls^7^. As an added benefit to the risk of phlebotomy, learners were offered a non-fasting lipid screening, measured by enzymatic reflectance spectrophotometry. This screening test is highly recommended for children and adolescents by the American Academy of Pediatrics but underutilized^8^.

### Regional Incidence of COVID-19

OCHCA collaborators routinely collect COVID-19 case rates across the county. To better understand the specific regional impact of COVID-19 at each of the four schools, we collated the countywide COVID-19 case data according to the zip codes of the learners at each of the schools during the two testing cycles (Figure 1).

**FIGURE 1.**
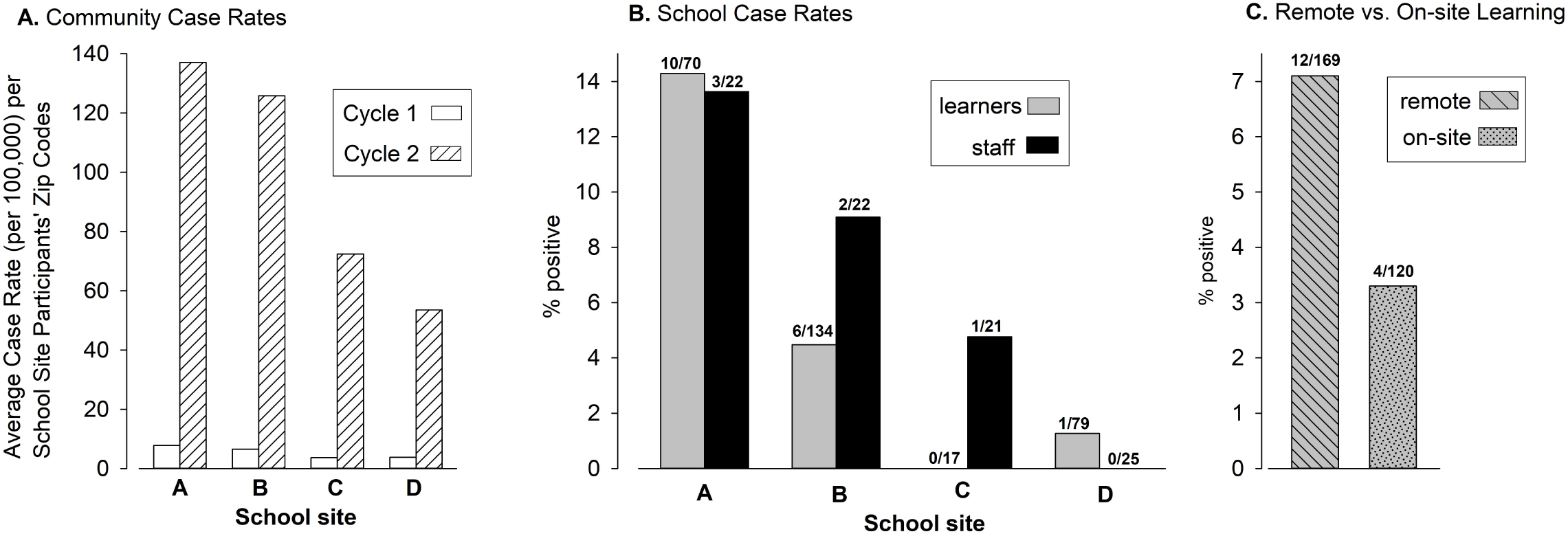
Regional and school-based SARS-CoV-2 positivity. (A) The marked growths in regional case rates (by participating students’ zip codes) are shown for each school site at testing cycle 1 and 2. (B) SARS-CoV-2 positivity by RT-qPCR in learners and staff at each of the school sites during cycle 2. (C) SARS-CoV-2 positivity by RT-qPCR in remote vs. on-site learners (aggregate of all 4 schools).

### Systematic Observation of COVID-19 Mitigation (SOCOM)

We adapted existing observation instruments, such as the System for Observing Play and Leisure Activity in Youth (SOPLAY) and the System for Observing Play and Recreation in Communities (SOPARC)^9,10^, to quantify the fidelity of face covering and physical distancing (≥ 6 ft.) mitigation in schools^11^. This new observation technique, Systematic Observation of COVID-19 Mitigation (SOCOM), used momentary time sampling techniques in which systematic and periodic scans of individuals were made in different (pre-determined) school environments (e.g., classroom, communal dining) and during physical activity (e.g., recess, physical education classes). Trained observers visited each of the 4 schools 3-5 times over a one-week interval and quantified mitigation in classrooms, recess, communal dining, and physical education (PE) classes.

### Statistical Analysis

All serology measurements and symptom ratings were treated as binary (yes/no) across the two visits. Lipid measures were each classified as normal or abnormal based on age-appropriate criteria. In the case of high-density lipoprotein (HDL) and low-density lipoprotein (LDL), the abnormal cases were further distinguished as low or high. Site comparisons of proportions, as well as cross-tabulations of two factors, were performed with Chi-Square analysis utilizing the Mantel-Haenszel correction. A α-level of 0.05 was used as the criterion for statistical significance. For learners found to be infected, age- and sex-matched uninfected peers were selected as controls for analysis. Immunological datasets were first tested for normality. To compare differences in various immune cell subsets between infected and age- and sex-matched uninfected participants, we used one-way ANOVA and Holm Sidak’s multiple comparisons tests. Group comparisons were tested using an unpaired t-test (Mann-Whitney U-test). For focus reduction neutralization assays, the half maximum inhibitory concentration (IC50) was calculated by non-linear regression analysis using normalized counted foci; 100% of infectivity was obtained by normalizing the number of foci counted in the wells derived from the cells infected with SARS-CoV-2 virus in the absence of plasma. Pearson correlation analyses were done by log transforming antibody end-point titers or neutralization titers.

## RESULTS

### Incidence of SARS-CoV-2 and Other Respiratory Viral Infections

No positive nasal RT-qPCR tests were identified in the first cycle of testing. During the second cycle, a total of 17 SARS-CoV-2 RT-qPCR positive results were observed among the 300 learners that were tested [5.7%, mean age 10.4 ± 2.1 (sd); range 7-17 y.o.]. Examination of SARS-CoV-2 community case rates by school site participants’ zip codes revealed low rates during the first cycle of testing, and a substantial rise in rates during the second cycle of testing (Figure 1A). As shown in Figure 1B, school A had the highest number of SARS-CoV-2 infected learners (p<0.01). In the aggregate (Figure 1C), there was no statistically significant difference in SARS-CoV-2 positive rates among remote or on-site learners (p=.1468). None of the hybrid learners (n=3) were positive for SARS-CoV-2. There was one additional case of SARS-CoV-2 among learners (n=8), who declined to disclose their education modality (i.e., remote, on-site or hybrid). In school B (the same geographic location as school A), we found 2 of 45 on-site learners (4.4%) and 4 of 89 remote learners (4.5%) had positive SARS-CoV-2 RT-qPCR. School D had 1 (1.3%) of on-site learners vs. none of remote learners had positive SARS-CoV-2 RT-qPCR. The low number of on-site participants in school A (n=2) prevented us from directly comparing SARS-CoV-2 positivity between remote and on-site instruction at each school site. Among the 90 staff and teachers tested in the second cycle visit, 6 (6.7%) were SARS-CoV-2 positive (Figure 1B). As with the learners, there were no positive results among the staff in the first cycle of testing. When normalized to data obtained from OCHCA zip-code-based case rates, school A showed the highest ratio of learner-to-local SARS-CoV-2 RT-qPCR positivity. We found that a significant number of learners in schools A and B had either low HDL or high LDL. In addition, 26% of learners with low HDL (p<0.0001) were also found to be SARS-CoV-2 RT-qPCR positive. There was no evidence of either influenza or RSV infection. Rhinovirus/enterovirus was observed in learners at all 4 schools (A: 9 of 72, 12.5%; B:15 of 142, 10.6%; C: 1 of 17, 5.9%; and D: 8 of 89, 9.0%). There was no correlation between SARS-CoV-2 and rhinovirus/enterovirus infections (p=.8397).

### Assessing Humoral and Cellular Immunity to SARS-CoV-2 in Children

Substantial numbers of learners (n=28) were found to have anti-NP IgG antibodies, indicating previous infection with SARS-CoV-2 (Figure 2A). As with the SARS-CoV-2 RT-qPCR results, there were large differences among the schools, most likely reflecting the differences in neighborhood infection rates (Figure 1A). There was also a significant association (p<0.0269) between SARS-CoV-2 RT-qPCR and anti-NP IgG results. Infected learners had detectable IgM and IgG titers against both the NP and the RBD of the spike protein (Supplemental Figure 1A), as well as neutralizing antibodies specifically to SARS-CoV-2 (Figure 2B, C). Neutralizing and binding antibody titers showed significant correlations (Supplemental Figure 1B). Moreover, learners with a history of SARS-CoV-2 infection generated broad and robust T cell responses as measured by IFN-γ ELISPOT following stimulation with overlapping peptides covering the entire viral proteome (Figure 3A). While the frequency of total CD4+ T cells was significantly lower in infected children (Figure 3B), the subset of proliferating (Ki-67+) CD4+ T cells was increased. This was driven by an increased proliferation within the effector memory CD4+ T cell (CD4+CD45RA-CCR7-Ki-67+) subset (Figure 3C). Levels of programmed death cell protein 1 (PD-1) were increased on CD4+ and CD8+ T cells from infected children indicative of recent activation (Figure 3D). Frequencies of circulating monocytes and natural killer (NK) cell subsets were lower in infected children (Figure 4A). There was no difference in inflammatory mediator responses (such as IL-1, IL-6, TNF-α) between healthy and SARS-CoV-2 seropositive children. Levels of innate immune cell activation markers such as HLA-DR were increased on myeloid dendritic cells (mDCs), while levels of FcγIII (CD16) and co-stimulatory molecule, CD86, were reduced on total NK cells and various monocytes subsets (Figure 4B, C).

**FIGURE 2.**
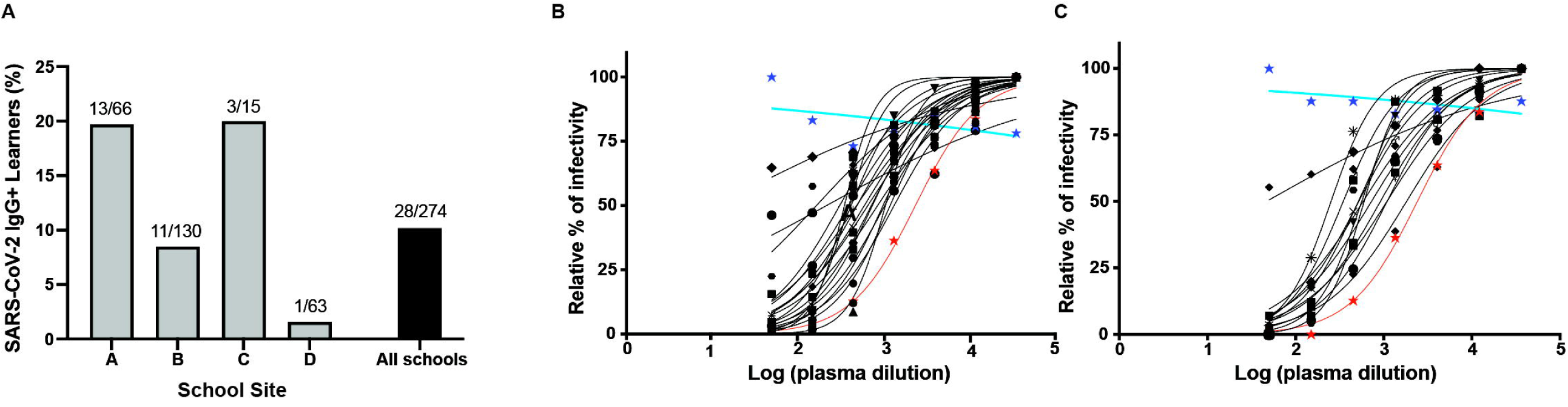
SARS-CoV-2-specific antibodies identified in school children. (A) Seroprevalence among learners at various schools. Neutralizing antibody titers against SARS-CoV-2 in learners from (B) testing cycle 1 and (C) cycle 2. Each learner is represented by a separate symbol and a best fit curve characterizing the neutralizing antibody capacity. The red line represents the positive control while the blue line represents the negative control. Infected, minimally symptomatic learners showed robust neutralizing antibody function to SARS-CoV-2.

**FIGURE 3.**
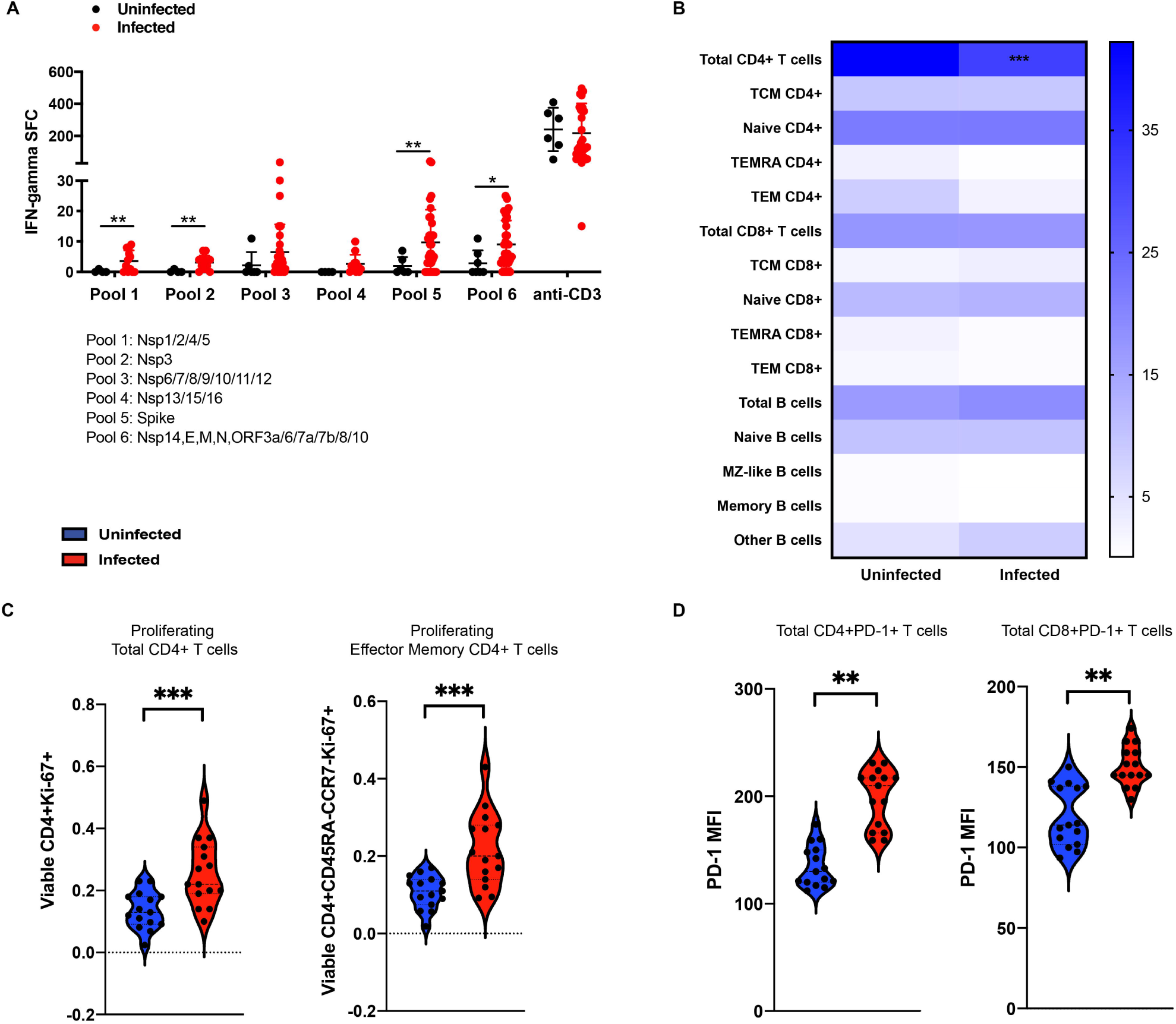
Effect of SARS-CoV-2 infection on adaptive immunity. (A) SARS-CoV-2-specific T cells were detected in seropositive learners by IFN-γ ELISpot assay following stimulation with overlapping peptides covering the entire viral proteome. (B) The frequency of adaptive immune cell subsets was identified by flow cytometry and revealed that the frequency of total CD4+ T cells was significantly lower in infected children. (C) There was nonetheless an increase in the proliferation of total CD4+ T cells (CD4+Ki-67+) and their effector memory subset (CD4+CD45RA-CCR7-Ki-67+). (D) Expression of programmed cell death protein 1 (PD-1), a marker of recent activation, on T cells of infected learners was also increased. Infected, minimally symptomatic learners showed robust adaptive immune responses to SARS-CoV-2. *p<0.05, **p<0.01, ***p<0.001.

**FIGURE 4.**
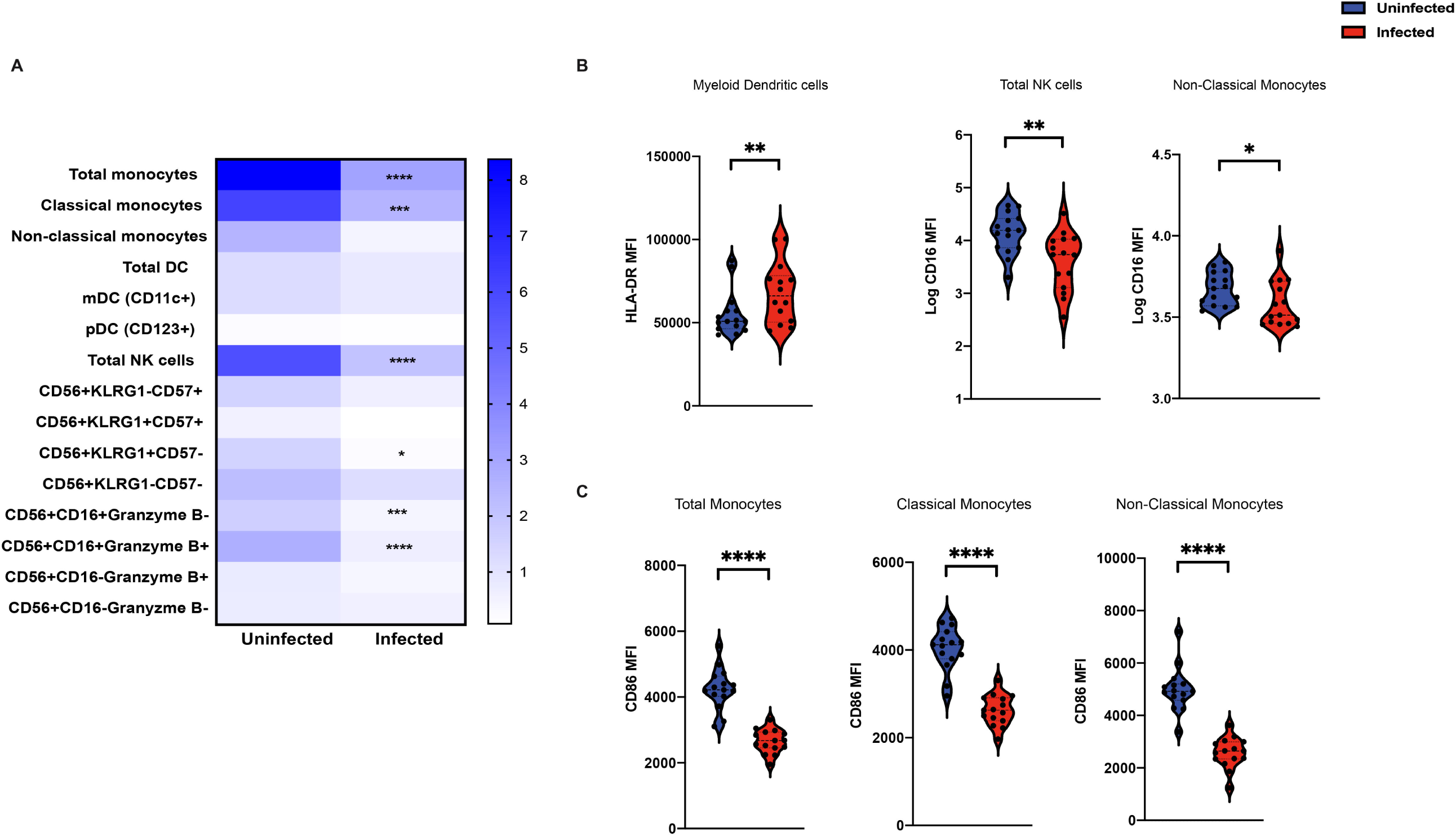
Effect of SARS-CoV-2 infection on innate immune cells. (A) The frequency of innate immune cell subsets was identified by flow cytometry and demonstrated decreased frequencies of circulating monocytes and natural killer (NK) cells in infected children. (B) The mean fluorescence intensity (MFI) of innate immune cell activation markers such as HLA-DR on myeloid dendritic cells (mDCs) was increased, whereas the expression of FcγIII receptor (CD16) on total NK cells and non-classical monocytes was reduced. (C) The expression of co-stimulatory molecule, CD86, on total monocytes, classical monocytes and non-classical monocytes was also reduced. *p<0.05, **p<0.01, ***p<0.001, ****p<0.0001.

### COVID-19 Symptoms

School A had the highest number of learners who reported symptoms associated with COVID-19 at 30% (p=0.0452). Rates for schools B, C, and D were 14.9%, 7.7%, and 18.6%, respectively. Learners who reported symptoms were significantly more likely to have SARS-CoV-2 positivity (13.6% vs. 3.5%, p<0.0018). Conversely, learners who were SARS-CoV-2 positive were more likely to have symptoms (47.1% vs. 16.8%, p=0.0018); in this group cough and fatigue were the most common.

### Systematic Observation of COVID-19 Mitigation (SOCOM)

SOCOM observations revealed high levels of face coverings and physical distancing compliance in classrooms at all four schools (Figure 5) and quantified the intuitively expected reduction in face coverings during communal dining (p<0.0001). At school D, which had the vast majority of on-site learners, face covering was consistently high (classroom, 96.7%; active recess, 97.3%; and PE, 97.0%). Physical distancing at school D varied (classroom, 81.6%; active recess, 35.1%; PE, 50.7%).

**FIGURE 5.**
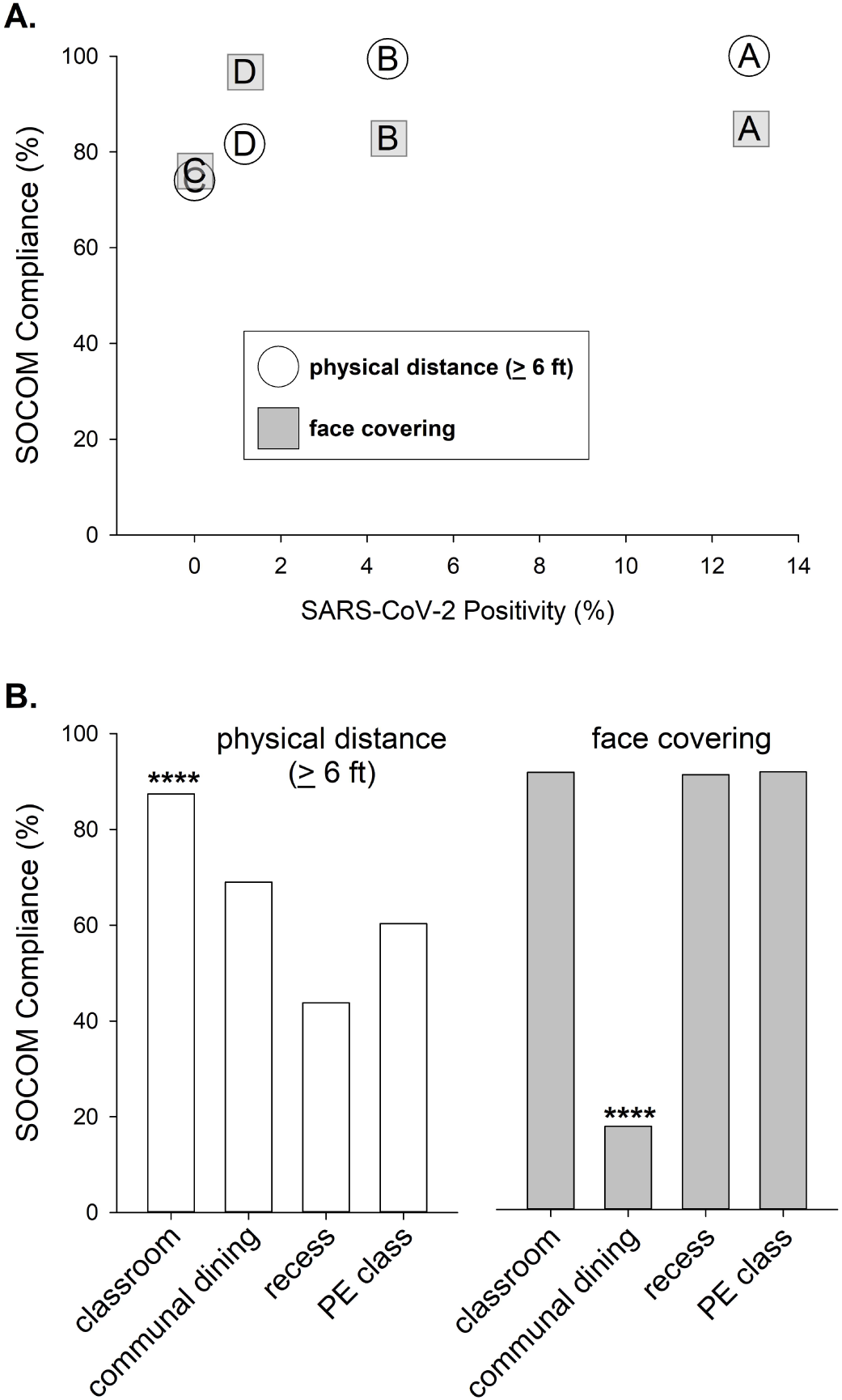
SOCOM data. (A) Physical-distancing and face-covering compliance was high in all schools and there was no correlation between in-classroom data and SARS-CoV-2 positivity. (B) Mean SOCOM values for classroom, communal dining, recess, and physical activity. Classroom physical-distancing was significantly highest, while face covering was lowest in communal dining. ****p<0.0001.

## DISCUSSION

This is one of the first school-based studies completed during the pandemic that directly and prospectively observed SARS-CoV-2 infection rates from school learners and staff. The four participating schools reflected the enormous diversity of income, community COVID-19 case rates, school type (private, charter, public), and learning status (remote vs. on-site) that face learners, school staff, and policy makers across the U.S. The huge increase in COVID-19 case rates between the two testing cycles permitted insight into the effect of the fall surge in SARS-CoV-2 infection. A unique feature of this research was that in contrast to the bulk of the data recently published on school-related SARS-CoV-2 infection, data was collected at the school site directly and not from data measured by report through public health agencies. Weaknesses of the study include 1) the possibility of selection bias as each participant and authorized legal guardian consented through a lengthy process in a time of great stress and anxiety, and 2) only one of our schools (school D) had predominately on-site learning.

Our results are consistent with several surveillance-based studies focused on schools during the pandemic. Zimmerman et al.^12^ implemented a comprehensive education and collaboration program and collected public health data in North Carolina; Falk et al.^13^ studied a rural Wisconsin community; and Macartney et al.^14^ studied preschools and K-12 schools in Australia. All studies concluded that secondary transmission of SARS-CoV-2 within schools was limited. We also found that infection rates reflected those of the community and neither remote learning nor highly mitigated on-site school attendance could eliminate SARS-CoV-2 infection. Comparisons and conclusions among our study and others done to date must be made with caution. For example, Zimmerman and Falk relied on public health agency contact tracing data. SARS-CoV-2 testing in learners or teachers would most likely result from a family reporting either symptoms or a known exposure to a primary care provider or testing center. Positive results would subsequently be linked to a particular school. Some infected individuals who were asymptomatic or symptom-deniers may not have been identified.

We show that under certain conditions, schools could host on-site learning with relatively low SARS-CoV-2 infection rates. The private school in our study (school D) remained open with a majority of on-site learners from July through December 2020, with few SARS-CoV-2 cases and low IgG positivity despite a tenfold increase in regional case rates. School D prepared for on-site learning by creating an advisory committee consisting of parents, local physicians, and content experts, and made an initial investment of about $1,400 per student to meet mitigation guidelines. While cost comparisons among schools serving very heterogeneous populations are always challenging, Rice et al.^15^ estimated that a comprehensive program of mitigation at schools would cost up to $442 per student.

Although SARS-CoV-2 infected children tend to report fewer symptoms than adults^16^, we found that school A had both the highest percentage of learners with symptoms and highest percentage of SARS-CoV-2 positivity. In addition, there was a significant relationship between SARS-CoV-2 positivity and presence of symptoms. These data support the use of limited symptom screening as a mechanism to enhance healthy school reopening.

We found that 26% of infected learners had significantly low circulating levels of HDL. Factors relating to obesity and physical activity are known to affect COVID-19 disease severity in adults and children^17–19^. Overweight and obesity are associated with these lipid abnormalities, all of which tend to occur with greater incidence in low-income school-aged children^20,21^. Levels of HDL seem to be particularly sensitive to physical activity^22,23^. The mechanisms responsible for the significant association that we found between low HDL levels and SARS-CoV-2 are at present unknown, but may be related to the association of overweight/obesity and chronic inflammation in children^24^. Increases in physical inactivity and weight in children have accompanied school closures over the past year^25,26^.

Widespread implementation of pediatric COVID-19 vaccination^27^ is many months away, and it is likely that adherence to COVID-19 mitigation procedures, including physical distancing and face covering, will need to continue for the near future. Previous studies cited above all highlighted the need to achieve high fidelity of COVID-19 mitigation procedures if viral transmission were to be limited. The in-classroom SOCOM data that we collected also revealed high fidelity at all four schools, including school C, which served many children with special needs, presenting additional challenges to COVID-19 mitigation. The successful implementation of mitigation procedures both in on-site settings and in the instruction to remote learners might have played a role in the complete absence of influenza virus that we observed^28,29^. In contrast, rhinovirus (which has the highest detection rate on school room desks among any respiratory viruses^30^) was observed in all schools. Implementation of quantifiable non-intrusive instruments like SOCOM along with testing of several respiratory viruses could help schools implement actionable strategies to limit SARS-CoV-2 transmission. The SOCOM method can help schools not only implement but also determine compliance with mitigation strategies.

Our immunological analyses revealed patterns that can explain the mild symptomatology that accompanies SARS-CoV-2 infection in most children. Frequencies of circulating total and classical monocytes, and expression levels of monocyte activation markers were lower in the infected compared with uninfected children. Moreover, key inflammatory mediators (e.g., IL-1, IL-6, and TNF-α) did not differ between the infected and uninfected comparison groups. This is in contrast to the monocytosis and heightened systemic inflammation observed in adult patients^31,32^. The absence of a heightened inflammatory profile, however, did not indicate a weaker immune response to SARS-CoV-2. Infected children generated robust and broad humoral and cellular immune responses and had detectable levels of SARS-CoV-2-specific IFN-γ secreting CD4+T cells following exposure to SARS-CoV-2 antigens. The infected children also had increased expression of PD1 on both total CD4+ and CD8+ T cells and a higher frequency of proliferating effector memory CD4+ T cells indicative of a recent history of activation.

The frequency of circulating cytolytic NK cells, those that mediate antibody-dependent cell cytotoxicity, was lower in the infected children. This observation corroborates previous studies in both children and adults, and supports the speculation that NK cells may be recruited into the lung^31,33–35^. Similarly, the frequency of circulating CD4+ T cells was reduced, suggestive of potential recruitment into the site of infection. These results support a maturation-dependent immune response to SARS-CoV-2 infection in children, one that specifically leads to milder disease and, possibly, to reduced transmission. The immune dysregulation that occurs in the rare but serious pediatric multisystem inflammatory syndrome in children (MIS-C) remains poorly understood^36^. However, based on the literature, MIS-C patients have much higher levels of circulating inflammatory mediators^37^ than what we observed in our study in acutely infected children who were asymptomatic to mildly symptomatic.

## CONCLUSION

This study indicated that neither remote nor on-site learning strategies could eliminate SARS-CoV-2 infection in school-aged children. Varied levels of successful infection prevention were observed in the four diverse schools studied that had differences in income level and regional levels of COVID-19 infection. The key challenge, of course, is balancing the damaging effects of school closures, which in the U.S. and throughout the world have adversely impacted low-income school-aged children and those with disabilities^38,39^, with the consequences of SARS-CoV-2 transmission to other learners and school staff. In retrospect, a larger, arguably national, more comprehensive approach to prospectively collecting SARS-CoV-2 infection patterns in school-aged children, their school staff and faculty, and family contacts would likely have provided the necessary information to achieve the shared goal of the healthiest environment for the continued education and physical and mental health of children and adolescents throughout the country. Our data do support the notion embodied in the Centers for Disease Control and Prevention (CDC) School Health Index that schools can effectively promote good health in children ^40^. We speculate that even at times of high community SARS-CoV-2 prevalence, schools can be among the healthiest places for children to be so long as the right mitigation strategies are in place. Finally, we would be remiss in not highlighting the remarkable dedication of the faculty and staff at all four schools who worked tirelessly to continue to provide meaningful learning to their students, and willingly and enthusiastically permitted us to intrude into their sites during an anxiety-provoking and uncertain time.

## Data Availability

The data that support the findings of this study are openly available in medRxiv.

## Abbreviations

CHOC: Children’s Hospital of Orange County
OCHCA: Orange County Health Care Agency
SOCOM: Systematic Observation of COVID-19 Mitigation
HDL: high-density lipoprotein
LDL: low-density lipoprotein
PBMCs: peripheral blood mononuclear cells
NP: nucleocapsid protein
RBD: receptor binding domain
NK: natural killer cells
mDCs: myeloid dendritic cells
PD-1: programmed death cell protein 1
MFI: mean fluorescence intensity

## ACKNOWLEDGEMENTS

The authors appreciated the outstanding management of this complex study by Phuong Dao, Brent Dethlefs, and the staff of the CHOC Research Institute.

## SUPPLEMENTAL MATERIAL FIGURE LEGENDS

**SUPPLEMENTAL FIGURE 1.**
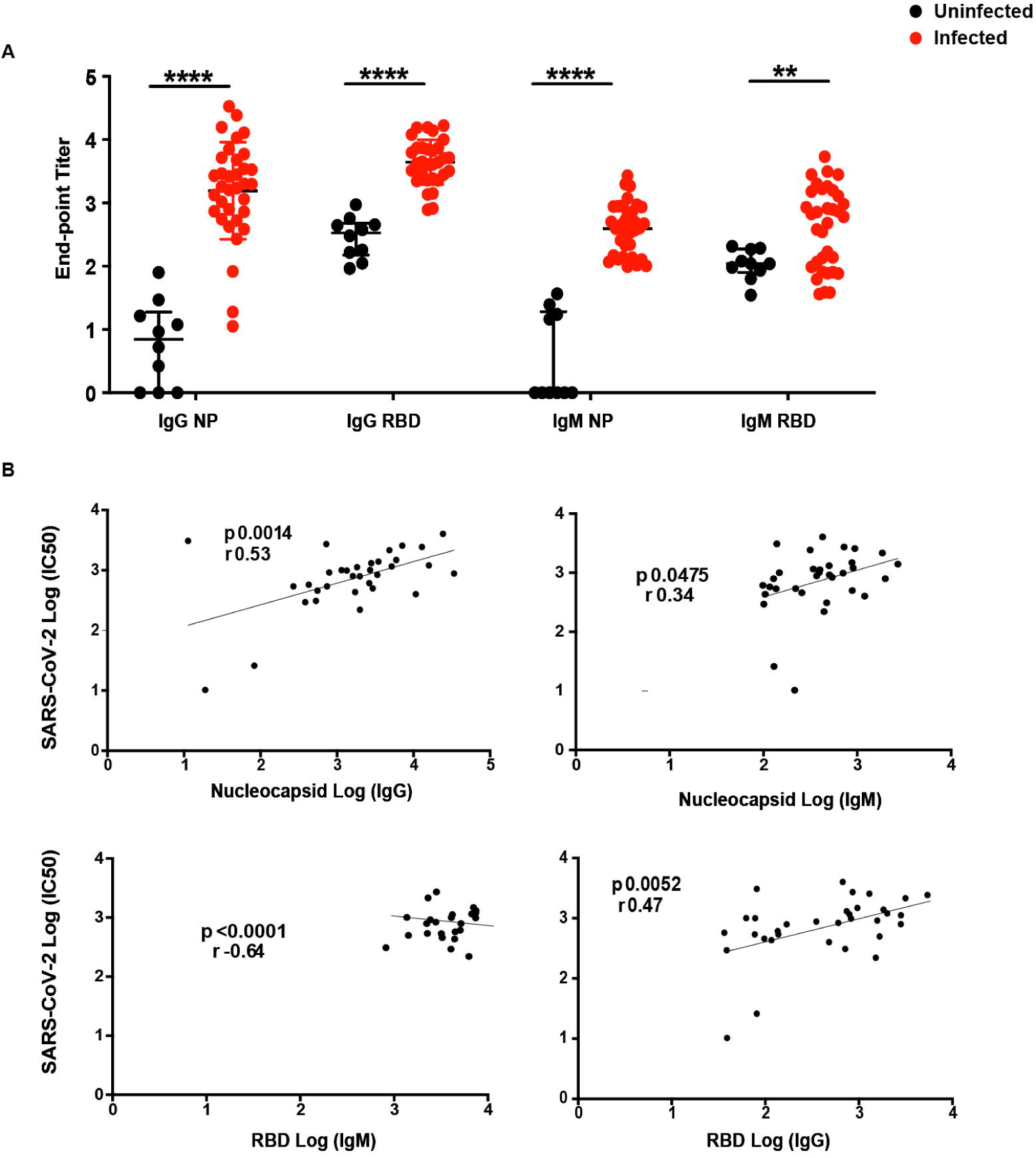
SARS-CoV-2 antibodies. (A) Detection of IgM and IgG end-point titers against SARS-CoV-2 nucleoprotein (NP) and receptor-binding domain (RBD) in learners. (B) Correlation between the half maximum inhibitory concentration (IC50) of the focus reduction neutralization titer (FRNT50) and IgM/IgG end-point titers in seropositive learners. **p<0.01, ****p<0.0001.

